# Molecular methods for diagnosis of monkeypox: A mini-review

**DOI:** 10.1101/2022.12.04.22283083

**Authors:** Rodrigo Michelini de Oliveira Thomasi, Thais da Silva Correa, Dalise Silva do Carmo, Déborah Fernandes Rodrigues, Luiz Vinicius da Silva Correa, Sandra Rodrigues Xavier, Liria Souza Silva, Jonatas Oliveira da Silva, Michelli dos Santos, Alessandra da Silva Dantas, Mariana Campos da Paz, Miguel Angel Chávez Fumagalli, Rodolfo Cordeiro Giunchetti, Eduardo Antônio Ferraz Coelho, Juliana Martins Machado, Alexsandro Sobreira Galdino

## Abstract

**Background:** Monkeypox is a global public health issue caused by the monkeypox virus (MPXV), a virus belonging to the Orthopoxvirus genus. As of October 28 2022, a total of 77,115 laboratory-confirmed cases and 3,610 probable cases, including 36 deaths, were reported, with 9,070 cases reported in Brazil, the second most affected country. The need to develop national technologies for the rapid diagnosis of emerging diseases for mass testing of the population is evident, as observed in the current SARS-CoV-2 pandemic. With that in mind, this article provides an overview of current methods, techniques, and their applications in the molecular detection of monkeypox.

**Methods:** The relevant documents or papers covered in this study were selected by a search in international bibliographic databases. The search terms used in the databases were aimed at summarizing existing knowledge on molecular diagnostic methods, such as: monkeypox; MPX, MPXV, qPCR, PCR, PCR-ELISA, and Diagnosis and Detection searched separately or together using the Boolean operator “AND” either in the title or abstract. The searches took place in September 2022, and the corresponding articles were selected between 2012 and 2022.

**Results:** We found 256 documents in total and twelve studies addressing the molecular diagnosis of monkeypox were classified as possible sources for this review.

**Conclusion:** This paper presents new perspectives and an overview of current methods, technologies, and applications in the molecular diagnosis of monkeypox. It is evident there is a pressing need to develop national technologies for the rapid diagnosis of emerging diseases for mass testing of the population. It is extremely important to have national detection kits with greater diagnostic capacity to assist in developing effective public policies in countries affected by this disease.

## INTRODUCTION

Monkeypox (MPX) is a zoonotic disease caused by the monkeypox virus (MPXV), an enveloped double-stranded DNA virus from the Poxvíridae family, Chordopoxvirinae subfamily, and the Orthopoxvirus genus (BUNGE *et al*., 2022; GONG *et al*., 2022; MORAND, A.; DELAIGUE, S.; MORAND, J., 2017). The NCBI Genbank (CLARK *et al*., 2016) was scanned and there are 2,695 genome assembles available for MPXV, which has a median total length (Mb) of 0.197488, median protein count of 179, and median GC% (guanine-cytosine percentage) of 33 (NCBI, 1982). MPXV was first discovered in 1958 among captive non-human primates in a Danish laboratory (MAGNUS *et al*., 1959). Although the virus was isolated for the first time in monkeys, its hosts also include rope squirrels, tree squirrels, Gambian rats, and voles (RIZK *et al*., 2022), and the natural cycle of the virus in nature is not fully known (ALTINDIS; PUCA; SHAPO, 2022).

MPXV can be divided into two different genetic groups: the Central African (Congo Basin) clade and the West African clade. Experts in pox virology, evolutionary biology, and representatives of research institutes around the world have reached a consensus to now refer to the former Congo Basin clade as clade one (I) and the former West African clade as clade two (II), which consists of two subclades. Clade I has been considered more transmissible and causes more severe diseases (NAKAZAWA *et al*., 2013; MARTÍN-DELGADO, M. C. et al., 2022; WILSON *et al*., 2014).

Nonetheless, monkeypox is a global public health issue, not only affecting countries in West and Central Africa, but also the rest of the world (KMIEC; KIRCHHOFF, 2022). In 1970, the first outbreak of monkeypox was reported in the Democratic Republic of the Congo (DRC) (LADNYJ; ZIEGLER; KIMA, 1972). The United States reported an outbreak of monkeypox in 2003, caused by contact with infected rodents (REED *et al*., 2004), and one of the biggest West African clade outbreak occurred in Nigeria in September 2017 (YINKA-OGUNLEYE *et al*., 2019). Nigeria has experienced a major epidemic with more than 500 suspected and more than 200 confirmed cases, with a fatality ratio of about 3% (WHO, 2022a). The current monkeypox outbreak, which is the largest one described so far in non-endemic countries, is thought to have started in the UK on May 6, 2022 with a British citizen who traveled to Nigeria where the disease is endemic, contracted the disease, and returned to the UK. As of May 21st, 92 cases have been confirmed in countries where the disease was not endemic (HRAIB *et al*., 2022; ISIDRO *et al*., 2022).

The mortality rate of monkeypox varies from 1 to 11%, which may depend on the clade. Until now, most reported deaths have involved young children and people with HIV who have not been vaccinated against smallpox (ADLER *et al*., 2022; DURSKI *et al*., 2019). The United States of America, Spain, Germany, Brazil, the United Kingdom, France, Canada, and The Netherlands present the highest number of reported cases. The outbreak clusters across Europe, the Americas, the Eastern Mediterranean, and Western Pacific regions are growing significantly (HEMATI *et al*., 2022). As of October 28 2022, a total of 77,115 laboratory confirmed cases and 3,610 probable cases, including 36 deaths, have been reported, with 9,070 cases in Brazil, the second most affected country (WHO, 2022b).

MPXV transmission occurs through large respiratory droplets, contact with skin lesions (close or directly), and likely through contaminated fomites. Sexual transmission through seminal or vaginal fluids is still inconclusive, but vertical transmission and fetal deaths have been documented (THORNHILL *et al*., 2022).

Sensitivity to risk factors and education on measures aimed at reducing exposure to viruses are the leading strategies for preventing monkey fever. Currently, no Food and Drug Administration (FDA) approved treatments for human monkeypox are available. However, research is currently underway to assess the feasibility and effectiveness of vaccines designed to prevent and control it. All the same, the smallpox vaccine, cidofovir, ST-246, and vaccinia immune globulin (VIG) can be used to combat monkeypox outbreaks. Even if these approved vaccines are not specific to MPXV, they could be prescribed for immunocompromised patients and in severe cases due to their effectiveness against smallpox (ALTINDIS; PUCA; SHAPO, 2022; SHAHEEN *et al*., 2022; WHO, 2022a).

The symptoms of monkeypox in humans are similar to those of smallpox but more severe. The incubation period lasts 7-14 days on the average but can also last up to 21 days. Initial symptoms include fever, headache, and muscle aches. A rash appears within 1-3 days after the onset of fever and tends to be more concentrated on the face and extremities rather than on the torso. Then, it spreads to other parts of the body (SHAHEEN *et al*., 2022; WHO, 2022a). Furthermore, one of the primary differences between monkeypox and smallpox symptoms is lymphadenopathy, which is present only in monkeypox (KUMAR *et al*., 2022).

Other rash illnesses might have similar clinical features to monkeypox, such as chickenpox, smallpox, measles, syphilis, bacterial infections, and allergies. Lymphadenopathy is an early sign that can be used to disguinsh monkeypox from chickenpox or smallpox. However, the absence of prodromal symptoms has been noticed in current outbreaks. Therefore, if it is clinically suspected, the confirmation of monkeypox is made through a polymerase chain reaction (PCR) test due to its accuracy and sensitivity. (ADLER *et al*., 2022; TARÍN-VICENTE *et al*., 2022; SOHEILI *et al*., 2022; WHO, 2022a). This paper provides new insights and an overview of current methods, techniques, and applications in the molecular diagnosis of monkeypox.

## 2. METHOD

A search in PubMed, Science Direct, Scielo, VHL, and Scopus databases resulted in the selection of the relevant documents or papers covered in this study. The search terms used in the databases were chosen to summarize existing knowledge on molecular diagnostic methods. The descriptors were: monkeypox; MPX, MPXV, qPCR, PCR, and PCR-ELISA. Diagnosis, and Detection were searched separately or together using the Boolean operator “AND” either in the title or abstract. The searches took place in September 2022 and the corresponding articles were selected between 2012 and 2022.

Regarding the inclusion criteria, electronically available articles in their entirety, addressing the topic in the title, abstract, or descriptors were selected. The exclusion criteria included: letters to the editor, case reports, editorials, duplicated articles, articles published in other languages, and those not directly dealing with the topic. Only studies addressing the molecular diagnosis of monkeypox were classified as possible sources for this review.

When a title or abstract appeared to describe a study eligible for inclusion, the articles were read in their entirety to assess their relevance as a way to reduce the risk of bias in individual studies.

## 3. RESULTS

This study analyzed documents from three molecular approaches: real-time polymerase chain reaction (qPCR), polymerase chain reaction (PCR), and polymerase chain reaction-enzyme linked immunosorbent assay (PCR-ELISA). The search for the PCR method resulted in 58 documents using (PCR monkeypox detection) AND (PCR monkeypox diagnosis) terms and Boolean operators. In the end, 55 papers were excluded and three were selected to report the PCR method for detecting MPXV. Also, the search for the PCR-ELISA method resulted in 10 articles using (PCR-ELISA monkeypox detection) AND (PCR-ELISAelisa monkeypox diagnosis) terms and Boolean operators. These documents reported the use of this technique but not in a combined way aiming at the enzyme-linked immunosorbent assay detection of the DNA. In this context, all papers were excluded. In addition, the qPCR method reported 178 documents using “qPCR OR real-time PCR” as terms. The terms “(Monkeypox OR MPX OR MPXV) AND (Detection OR diagnosis)” were also included along with the Boolean operators. Nine articles were selected according to our inclusion criteria. The search flowchart is shown in **Figure 01**.

## 4. DISCUSSION

### 4.1 Real-time Polymerase Chain Reaction

Currently considered the gold standard in the diagnosis of monkeypox, qPCR is an evolution of conventional PCR, differing from it in the detection of the amplified product. While in conventional PCR this step is performed by means of gel electrophoresis, in qPCR it is made through fluorescence detection, which is performed during the amplification process, giving the assay greater agility in obtaining the results with a lower risk of contamination and better accuracy (JALALI, ZABOROWSKA, and JALALI, 2017). Detection simultaneously with amplification can be done using fluorescent dyes that nonspecifically bind to double-stranded DNA or specific probes that bind to certain DNA sequences and contain a reporter fluorophore that emits fluorescence, indicating that amplification has taken place (TAJADINI, PANJEHPOUR, and JAVANMARD, 2014). Therefore, the fluorescence measurement is used to quantify the product of interest, which can be absolute, when the result is obtained by comparing the emitted fluorescence with a standard curve or relative, when the signal of the sequence of interest is compared with a control signal (PFAFFL, 2012).

The following studies were compiled in Table 1, containing the primers, probes, and terminal dye primers used for each study.

**Table 1.**
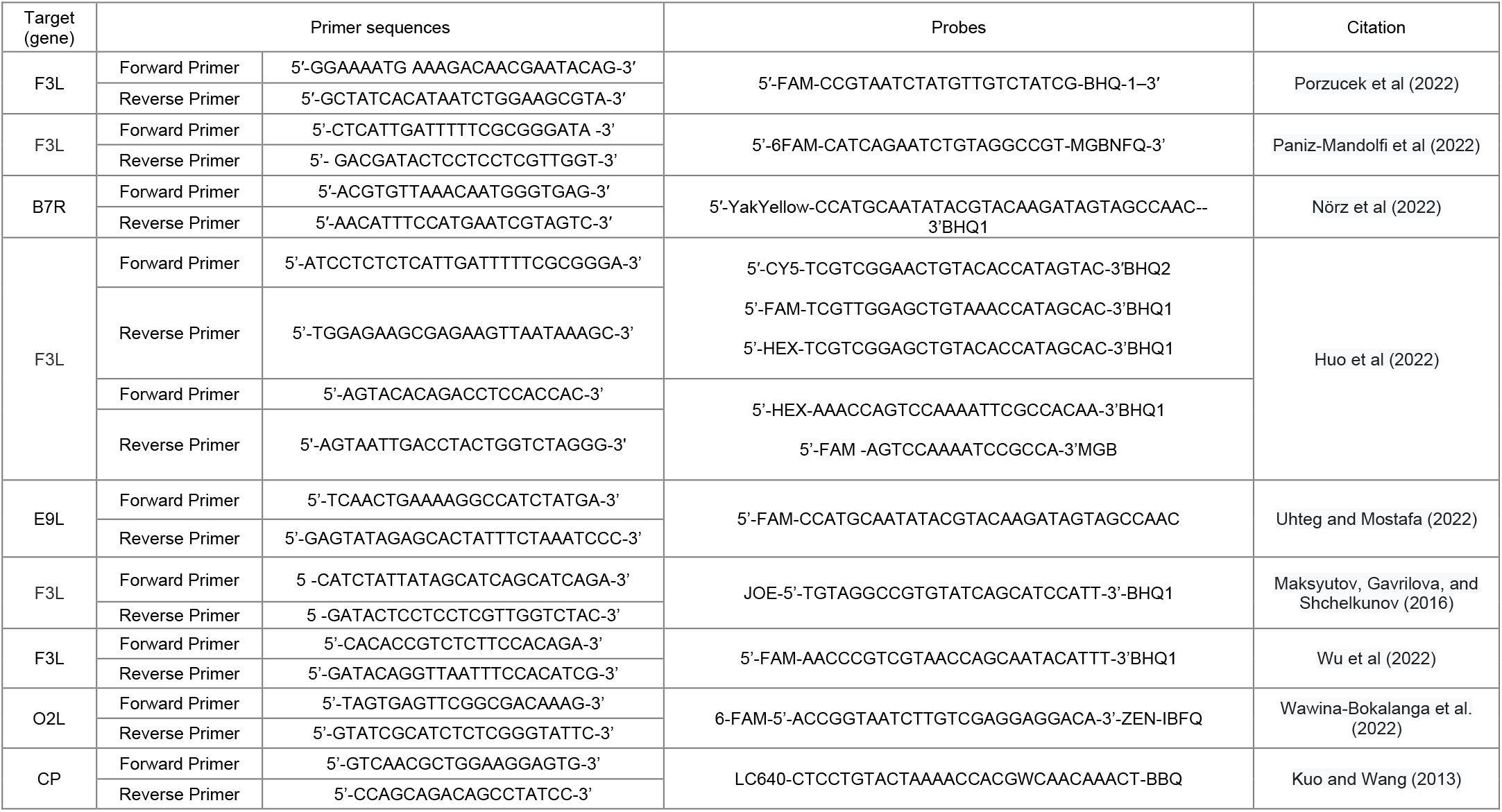
Primers sequence used in diagnostic research

Porzucek *et al*. (2022) developed and validated an accessible qPCR assay for MPXV detection. MPXV was detected with cycle threshold (Ct) values of 17.46–35.59, titer equivalents to 8.01×106 to 2.45×100 plaque–forming unit (PFU)/mL. A detection limit of 3.59 PFU/mL and sensitivity and specificity of 100% were reported.

Paniz-Mandolfi *et al*. (2022) evaluated and validated a novel, sensitive, and specific qPCR method for MPX diagnosis in humans. The specificity, sensitivity, and analytic performance were evaluated, in which the limit of detection was 7.2 genome copies/reaction. The virus was identified in twenty clinical specimens with 100% correlation against a reference method and sensitivity/specificity were both 100%.

Huo *et al*. (2022) proposed and evaluated two multiplex real-time PCR assays for the simultaneous detection and differentiation of MPXV with high specificity and sensitivity. The primers and probes that were used for Assays 1 and 2 are shown in Table 1. Specific probes based on four nucleotides were designed between MPXV-WA (IIa, IIb) and MPXV-CB (clade I), which were then mixed to quickly detect and differentiate the two clades. That way, a universal test with wide application possibilities would be created. Probe specificity was assessed by mixing a pair of primers and three probes in Assay 1 to test three types of samples. The IIa, IIb sequence only matched the FAM-labeled probe at the 5’ end, while the MPXV-I clade sequence was only efficiently annealed to the CY5-labeled probe at the 5’ end. No cross-reactivity was detected between MPXV-IIa, IIb, MPXV-I, and other orthopoxviruses, demonstrating that this assay has the high specificity. The amplification curves obtained by mixing the two probes were compared in Assay 2. There were amplification curves in two fluorescence channels when placed in the plasmid of the MPXV B.1 strain. MPXV B.1 could be identified by the TaqMan probe labeled with HEX at the 5’ end, while other strains did not cross-react with this probe.

Uhteg and Mostafa (2022) aimed at the validation and implementation of a qPCR assay for orthopoxvirus for clinical use in the diagnosis of MPXX, using the primers described in Table 1. To determine the specificity of the proposed test, samples from monkeypox-positive patients were also run in-house with a PCR-based search specific for monkeypox, using the primers and probes described in the literature, targeting the B6R gene. The results showed that the primers and probe selected for the assay target regions that are conserved in genomes associated with the 2022 outbreak. The assay showed sensitivity of 100 copies/ mL, with 100% reproducibility and specificity. In addition to establishing the stability of the samples for clinical testing after as long as a week of storage at room temperature, the assay demonstrated a positivity rate of 33.33% during the first month of use at clinical testing.

Maksyutov, Gavrilova, and Shchelkunov (2016) described a method for species-specific identification of human pathogenic variation and MPX with simultaneous varicella-zoster virus detection in a multiplex TaqMan qPCR assay. For this, the authors used oligonucleotide primers and corresponding hybridization probes for the region of the B12R gene for the smallpox virus, F3L gene for MPXV, and the ORF38 gene for varicella-zoster virus described in Table X: VARV_B12R_probe; VARV_B12R_superior; VARV_B12R_lower; MPXV_F3L_probe; MPXV_F3L_upper; MPXV_F3L_lower; VZV_ORF38_probe; VZV_ORF38_upper; VZV_ORF38_lower and IC_probe. The analytical specificity of the multiplex qPCR assays was equal to 100%. The reproducible detection limit was approximately 20 copies for VARV and MPXV and 50 copies for VZV DNAs per reaction, thus demonstrating the potential of this method to be used in the control of MPXV outbreaks.

Wu *et al*. (2022) analyzed the primer and probe sequences in the generic qPCR assay recommended by the MPXV Centers for Disease Control (CDC), as well as the specificity of seven qPCR assays aimed at detecting MPXV and other orthopoxviruses. An analysis of the primer and probe sequences was performed by aligning 683 MPXV genomes reported worldwide. Sequence mismatches were found in more than 92% of the genomes for the generic forward and reverse MPXV primers. Assessment of the specificity of the real-time PCR assays identified two assays (MPXV_F3L and OPV_F8L) with the highest match score (>99.4%) for the global MPXV genome database. Genetic variations in the MPXV genomes corresponding to the real-time PCR primer and probe regions were specified and used to indicate the temporal and spatial emergence pattern of monkeypox disease. The results showed that the current generic real-time MPXV assay may not be adequate to accurately detect MPXV, highlighting the need to develop new detection assays.

Wawina-Bokalanga *et al*. (2022) developed a rapid and highly efficient, MPXV-specific qPCR. Initial assay validation was performed on Mic qPCR and ABTM 7500 Fast Real-Time PCR systems. The performance was proved with five MPXV-positive DNA, and five out of ten clinical samples tested positive with Ct values ranging from 20.6 to 34.9. The primers were specific for all known MPXV genomes, and it can still detect viruses of both the Central and West Africa MPXV clades, including the clade that is currently circulating in Europe.

Kuo and Wang (2013) reported two pan-Orthopoxvirus (pan-OPV) SYBR® green-based PCR assays targeting late 16-Kda putative membrane protein (LPMP) and CP genes, with high sensitivity in detecting pan-OPV DNAs, including MPXV, in levels of 1 copy DNA/reaction or 10 pfu/ml. Monkeypox probe-based real-time PCR assays targeting 39-kDa core protein (CP) genes can sensitively and accurately detect monkeypox and variola at the level of 10 copies and 1 copy of DNA per reaction, respectively.

Nörz *et al*. (2022) adapted two qPCR assays (non-pox virus specific and pox virus specific) for use as a dual-target MPXV detection test. In this approach, one assay targeted a conserved sequence from the Orthopoxvirus genus, not including major or minor pox virus, and the other targeted a specific MPXV sequence. An in silico evaluation of potential primer-probe interactions was performed and the chosen primers and probes are described in Table 1. Submitted sequences were aligned to currently available MPXV and orthopoxvirus sequences available in public databases. The evaluation of the analytical performance of the proposed test showed a detection limit of 95%. Specificity was assessed by testing 53 specimens, including clinical specimens, reference material, and external quality controls for a variety of respiratory and bloodborne pathogens. There were no false positives or false negatives, showing 100% sensitivity and specificity.

### 4.2 PCR and PCR-ELISA

The polymerase chain reaction (PCR) is a technique widely used in molecular biology. At present, PCR represents the gold standard for the detection of viruses such as HPV (MENÊSES, TORALLES, MENDES, 2019) and cytomegalovirus (EXLER *et al*., 2019), Sars - Cov-19 (WANG *et al*., 2020), among others. The PCR technique consists of the artificial multiplication of the genetic material of interest in a regulated manner, and one of the most important factors of the technique is the use of primers that will be responsible for the enlarged DNA section. Since the first monkeypox outbreak occurred in 2003 in the United States, the PCR technique has been established in the analysis and molecular diagnosis of the viruses that cause the disease. Two types of assays were then established, the first capable of detecting the DNA polymerase gene in Eurasian orthopoxviruses and the second targeting an envelope protein (B6R), the B6R protein being specific to the monkeypox virus (LI, 2006). However, the focus of PCR analysis has changed in the wake of several other outbreaks over the last 20 years. It is evident that the use of PCR developed a more detailed profile, looking for origins and lineage differentiation. Li (2010) developed new assays for the identification of West African and Congo Basin clades using TaqMan probe technology.

As shown by Saijo (2008), the A-type inclusion body gene (ATI) can be used to differentiate between Congo basin clades and West African clades. Erez *et al*. (2019) reported the use of PCR for the diagnosis of the first case of monkeypox in Israel. In 2018, samples taken from a patient with clinical signs of the disease were analyzed at the Israel Institute for Biological Research. The PCR diagnosis was based on specific primers to discriminate between the West African (581 bp) and Congo-Basin (832 bp) clades by product size. The PCR found product size corresponded to that of the West African clade circulating in Nigeria at the time. Thus, the study group was able to successfully identify the first imported monkeypox case in Israel.

Kim *et al*. (2022) used PCR to detect MPXV from skin lesions, nasopharyngeal, and oropharyngeal swabs obtained from the first monkeypox case in the Republic of Korea. The following primers were used: F3L-forward primer, F3L-reverse primer; A39R-forward primer, A39R-reverse primer, 1995MPV-forward primer, 1995MPV-reverse primer, and 5’-TTGTATTTACGTGGGTG-3’ for the B2R. Conventional PCR showed a 1,550 pb product, which confirms the MPX diagnosis, indicating that the MPXV belonged to the West African clade.

Tumewu *et al*. (2020) related the use of PCR technique to perform a differential diagnosis of an adult patient suspected of monkeypox infection. A 51-year-old male patient presenting symptoms similar to monkeypox was admitted to a hospital in Surabaya, Indonesia. Vesicle fluid was collected for PCR amplification, which was performed with specific primers either for MPXV or the chicken pox virus (VZV). The PCR was followed by genome sequencing that showed homologies of more than 99 % to other VZVs and less than 50% to monkeypox sequences.

Despite the PCR’s efficiency, this technique still has some limitations, such as the need for a step to identify the amplified products. The detection step is usually performed using gel electrophoresis. This step represents a significant slice of the time required for diagnosis and a low detection threshold. Additionally, gel electrophoresis only allows the detection of the presence or absence of a particular gene (SUE *et al*., 2014). Gel electrophoresis also exposes the professionals who perform the test to health risks, not to mention damage to the environment, as it requires using the toxic reagent ethidium bromide (RAMI, AMANI, SALEH, 2019). Thus, the PCR technique has been modified over the years to minimize these limitations. Combinations with other established techniques have been increasingly used to enhance the sensitivity and specificity of the PCR diagnosis. Among these combinations, PCR-ELISA stands out, which emerged as a synergistic combination of two techniques, using the ELISA assay as an alternative to the electrophoresis step.

ELISA is a quantitative analytical method capable of identifying identify antigen-antibody reactions through a color change, which is obtained from an enzymatic conjugate and its substrate. The enzyme-substrate reaction is used to identify the presence and amount of molecules in biological fluids (HORNBECK, 1992). This method is called enzyme immunoassay. It emerged as an alternative to immunodetection tests that, until the 1960s, used radioactive markers. ELISA was developed in 1971 by Swiss researchers Engvall and Perlmann by modifying the radioimmunoassays used at the time. In the new method, radioactive iodine was replaced by enzymes (AYDIN, 2015). The method exploits the specific features of the antigen-antibody interaction, such as binding specificity through non-covalent interactions (MURO *et al*., 2009). Since then, this immunoassay has been widely used for various purposes (ENGVALL, 2010). To perform this assay, the antigen is fixed to a surface, with a 96-well plate being commonly used. An antibody bound to an enzyme tag is then exposed to the antigen attached to the well. The wells are then washed to remove all unbound compounds. The detection is completed when the presence of the marker is analyzed after washing the wells and adding a chromogenic substrate, which, after aeration, changes its color. Thus, when the antigen under analysis is present, the detection equipment perceives and quantifies the observed color change, whose intensity refers to the concentration of substrate that reacted to the enzyme, attesting to a greater or lesser concentration of the antigen (AYDIN, 2015).

The first studies on DNA immunomodulation began in the late 1980s (COUTLÉE *et al*., 1989). Since then, several studies have been developed in this area, culminating in the development of a method that combines these two techniques, the PCR-ELISA. It consists of a single analytical technique with a protocol similar to that of ELISA, only unlike the former, it detects nucleic acid instead of protein (DI PINTO *et al*., 2012). Since the DNA fragment is labeled with biotin or digoxigenin, it can be easily identified through a color change, similar to the detection of proteins through the enzyme antibody reaction. Thus, PCR-ELISA is able to quantify the PCR product directly after DNA immobilization in a microplate. It is performed in three steps: PCR amplification, immobilization of the amplified product on microplates, and the colorimetric detection of this product (SUE *et al*., 2014). PCR-ELISA allows a large number of samples to be processed in less time, using an environmentally friendly approach with high sensitivity (SAMIMI *et al*., 2022).

Despite being potentially effective in the molecular diagnosis of monkeypox, the PCR and PCR-ELISA techniques are not being used primarily in the detection of this disease. Although conventional PCR and PCR-ELISA techniques play a significant role in molecular diagnosis and may well represent a promising alternative for MPXV detection and MPX diagnosis, few articles in the selected time period report the use of conventional PCR as the only resource for diagnosis and detection of MPXV, and none of them report the use of PCR-ELISA. This is most likely due to the fact that the gold standard for MPX virus detection in this new outbreak is real-time PCR, despite the efficiency of the aforementioned techniques. Real-time PCR combines speed and sensitivity, resulting in a diagnosis with a very low detection limit.

### 4.3 Other molecular approaches

Other molecular methods were found during the literature search for conventional PCR, PCR-ELISA, and qPCR techniques. Feng *et al*. (2022) reported the use of loop-mediated isothermal amplification (LAMP), a variation of the conventional PCR using enzymes that allow amplification at constant temperatures, thus dispensing with the use of specialized equipment in the development of a specific assay for MPXV detection. The authors used primer sets A27L-1 and F3L-1 at 63°C. The detection limits of the LAMP assay were 20 copies/reaction mix, 0.100 times higher in sensitivity compared to conventional PCR. The high specificity of the assay was verified by testing 21 non-MPXV pathogens, with negative results for all pathogens tested, thus confirming the effectiveness of the proposed assay, which can provide faster on-site diagnosis, especially in primary hospitals and rural areas. Bhadra and Ellington (2022), in turn, reported the use of LAMP to amplify two regions of the MPXV genome using primers for the J2L genes (FIP, BIP, F3, B3, and one LP) and A26L. Detection of the presence of MPXV is made by fluorescence analysis, which, after replication at 60°C for 60 minutes, showed a strong increase in fluorescence, demonstrating the effectiveness of the test.

Another technique is the amplification of recombinase polymerase (RPA), a variation of isothermal amplification that uses strand transfer DNA polymerase, which allows the reaction to occur at a constant temperature, between 37 to 42°C or even at room temperature, albeit a little slower in this case. Davi *et al*. (2022) used this technique to develop a test capable of identifying MPXV clades from Central and West Africa targeting the G2R gene. The assay was performed at a constant temperature of 42°C, with a detection limit of 16 DNA molecules/μl and results after 3 to 10 minutes. Its specificity was 100%, while sensitivity was 95%.

Methods based on the Clustered Regularly Interspaced Short Palindromic Repeats (CRISPR) system have also stood out. In bacteria, the CRISPR-Cas system recognizes viruses that have previously infected the microorganism, producing RNA probes that will interact with the viral genetic material. These probes direct a Cas-like enzyme to the site, which cut the viral genetic material where it interacted with the probes (JACKSON *et al*., 2017). Sui *et al*. (2022) developed an MPXV detection system that uses DNA target analyte P4-WT (BHQ-GGGTTAGGGTTAGGGTTAGGG-FAM) labeled with 6-fluorescein (6-FAM). When binding to the viral DNA, the analyte undergoes the action of the Cas 12 enzyme, which cleaves the 5’ end, resulting in the emission of a fluorescent signal. The fluorescent signal was detectable in 2 min, with a strong emission after 10 min, which suggests that the developed system is a potentially fast detection technology.

Finally, genomic sequencing also represents a method that can be used to detect viral differentiation. Israeli *et al*. (2022) produced a rapid amplicon nanopore sequencing system (RANS), aiming at the differential diagnosis of MPX. The approach used by the authors presupposes the use of diagnostic regions of the viral genome that have high homology in their limits that, when sequenced, help identify the pathogen. To carry out the study, a dA tail and a 50-phosphonate were added in the multiplex PCR amplification step, thus preparing the PCR product for sequencing, performed in a nanopore system. The sequences were compared to databases, thus identifying the pathogen. After rapid sequencing (a few minutes), the readings were compared to a reference database and the closest strain was identified. The tests showed 100% sensitivity and specificity for the evaluated pathogens.

## 5. CONCLUSION

The threat of a monkeypox pandemic is real. Reports of new cases are spreading across most regions of the globe, demanding a quick response from government authorities. The current outbreak has once again highlighted the need to develop fast and efficient tests for the detection of MPXV and early diagnosis of monkeypox. In addition, molecular methods are among the safest and most accurate diagnostic methods, including the current gold standard for monkeypox diagnosis, qPCR. This technique has characteristics such as high sensitivity and specificity, in addition to delivering faster results than other molecular methods, such as conventional PCR and PCR-ELISA. As demonstrated, new methods have been developed to ensure the rapid diagnosis and effective monitoring, which are safe and fast, sometimes at a lower cost, providing crucial advances that allow a better response and management of the new outbreak of MPX, especially in developing countries.

This paper offers new perspectives and a general view of current methods, technologies, and applications in the molecular diagnosis of monkeypox, which are still insufficient due to a burgeoning demand. The current SARS-CoV-2 pandemic brought new lessons, one of them being the need to develop national technologies for the rapid detection of emerging diseases and mass testing of the population. It is vitally important to have national kits with a greater diagnostic capacity to create public policies in countries affected by this disease. In Brazil, we can cite two public policies in the field of biotech such the action plan for science, technology and innovation (PCTI Biotecnologia (2018-2022) and the Brazil Initiative (Ordinance No. 4,488, of February 23, 2021) that encourages the creation of companies based on technology as startups in Life Science for the development of innovative products as diagnostics kits.

## 6. CURRENT & FUTURE DEVELOPMENTS

A decline in smallpox immunity due to the cessation of vaccination in 1980 paved the way for the disease to resurge and spread to non-endemic parts of the world. There has been a significant rise in the number of individuals infected with smallpox, with the highest rates among men (EJAZ *et al*., 2022; BRAGAZZI *et al*., 2022; HARAPAN *et al*., 2022; KOZLOV, 2022; VIVANCOS *et al*., 2022). As a result, health care professionals must be on high alert and make sure their knowledge and skills are continously being updated (LUO; HAN, 2022; HARAPAN *et al*., 2022).

Studies have shown that gaps in knowledge about smallpox and low levels of confidence in diagnosing, managing, and preventing the disease are prevalent among health care professionals and medical students (HARAPAN *et al*. al., 2020a; b). In addition, neurologists and physicians should be aware of the possible neurological complications caused by MPXV (SEPEHRINEZHAD; ASHAYERI AHMADABAD; SAHAB-NEGAH, 2022).

PCR is the modality of choice for laboratory testing and has been widely used to detect MPXV (PETERSEN *et al*., 2019). Although sensitive and selective, PCR-based MPXV detection approaches are not suitable for resource-limited settings. The MPXV diagnostic development timeline shows there has been limited progress toward innovations in MPXV diagnostics, indicating a research gap, and new methods of detecting MPXV antigens should be developed in the near future. The COVID-19 pandemic has brought new lessons, one of them being the need to develop technologies for the rapid diagnosis of the population and implement effective public policies in countries reporting an outbreak of this disease since imported kits are costly, place a burden on the public health care system, and make it difficult to detect and control the disease. The expectation is for more new methods with a lower cost, greater reliability, and faster results (DE OLIVEIRA *et al*., 2022). Several preclinical studies have been conducted to find a potential cure for monkeypox and data on the safety and efficacy of the drugs are critical (HARAPAN *et al*., 2022).

There is insufficient evidence to determine whether oral brincidofovir (inhibitor of DNA polymerase), oral tecovirimat (inhibitor of intracellular viral release), or intravenous immunoglobulin vaccinia are effective against MPXV. To produce a possible therapeutic antiviral agent requires more in-depth studies at the genomic level and molecular analyses to better understand the virus-host interaction (HARAPAN *et al*., 2022).

All available smallpox vaccines offer good protection against infection and can be used as a pre-exposure or post-exposure prophylaxis (VOUGA *et al*., 2022); the two smallpox vaccines, JYNNEOS (Imvanex/Imvamune) and ACAM2000, provide 85% effectiveness in preventing the disease (EJAZ *et al*., 2022). Vaccination and measures such as family hygiene education and patient isolation will be crucial in the future control of monkeypox outbreaks and other emerging or re-emerging orthopoxviruses.

## Data Availability

All data produced in the present work are contained in the manuscript

## AUTHOR’S CONTRIBUTION

#Rodrigo Michelini de Oliveira Thomasi and Thais da Silva Correa contributed equally to this work

## CONFLICT OF INTEREST

The authors declare no conflict of interest, financial or otherwise.

## ACKNOWLEDGEMENTS

The authors would like to thank the Universidade Federal de São João Del-Rei (UFSJ), Fundação de Amparo à Pesquisa do Estado de Minas Gerais (FAPEMIG), Coordenação de Aperfeiçoamento de Pessoal de Nível Superior (CAPES), and Conselho Nacional de Desenvolvimento Científico e Tecnológico (CNPq) for all their support. ASG also thanks CNPq for its productivity fellowship DT.

## Notes

### Competing Interest Statement

The authors have declared no competing interest.

### Funding Statement

This study did not receive any funding

